# Online queries as a criterion for evaluation of the epidemiological status and effectiveness of COVID-19 epidemic control measures

**DOI:** 10.1101/2021.07.11.21260148

**Authors:** K. T. Momynaliev, D. K. Khoroshun, V. G. Akimkin

## Abstract

Monitoring online queries can provide early and accurate information about the spread of COVID-19 in the population and about the effectiveness of COVID-19 epidemic control measures.

**The purpose of the study:** Assessment of significance of online queries regarding smell impairment to evaluate the epidemiological status and effectiveness of COVID-19 epidemic control measures.

**Materials and methods:** Weekly online queries from Yandex Russian users regarding smell impairment were analysed in regions and large cities of Russia from 16/3/2020 to 21/2/2021. A total of 81 regions of Russia and several large cities, such as Moscow, St. Petersburg, and Nizhny Novgorod, were included in the study.

**Results:** A strong positive direct correlation (r>0.7) was found between the number of smell-related queries in Yandex new cases of COVID-19 in 59 out of 85 Russian regions and large cities (70%). During the “first” peak of COVID-19 incidence in Russia (April-May 2020), the increase in the number of smell-related queries outpaced the increase in the number of new cases by 1-2 weeks in 23 out of 59 regions of Russia. During the “second” peak of COVID-19 incidence in Russia (October-December 2020), the increase in the number of smell-related queries outpaced the increase in the number of new cases by 1-2 weeks in 36 regions of Russia, including Moscow. We also estimated the increase in the query/new case ratio during the “second” peak of incidence for 45 regions. It was found that the query/new case ratio increased by more than 100% in 24 regions. The regions where the increase in queries was more than 160% compared to new infection cases during the “second” peak of incidence demonstrated significantly higher search activity related to levofloxacin than the regions where the increase in queries was lower than 160% compared to the increase in new infection cases.

**Conclusion:** The sudden interest in smell impairment and growing frequency of online queries among the population can be used as an indicator of the spread of coronavirus infection among the population as well as for evaluation of the effectiveness of COVID-19 epidemic control measures.

## Introduction

In recent years, big data analytics have become increasingly integrated in studies conducted in public health informatics, and web data analysis has become a valuable tool for monitoring and population behaviour analysis in terms of health-related content. The term for using data from web-based sources to improve public health is known as “infodemiology”, a combination of the words “information” and “epidemiology” [1]. Infodemiology and infoveillance studies using different web-based sources, such as Google, Twitter, and other social media platforms, show the importance of real-time access to data when evaluating health status [2, 3, 4, 5, 6].

On the other hand, during the global COVID-19 pandemic, the scale of public interest in this disease has been unprecedented, suggesting that the trends demonstrated by web search traffic should remain steady and reliable. Changes in smell and taste are prominent symptoms of COVID-19 [8–12], as has consistently been demonstrated in many countries (e.g., Iran, Spain, France, Italy, Germany, and the UK) [8, 13-16]. More critically, these chemosensory changes generally occur earlier than other symptoms and may constitute more specific symptoms than fever or dry cough [8, 15, 17]. Accordingly, web-based monitoring of changes in smell and taste can provide early and specific information on the spread of COVID-19 in the general population and support health system monitoring to evaluate the effectiveness of epidemic control measures taken by countries against COVID-19.

In a previous study, we demonstrated a strong correlation between the relative search volume (RSV) in Russia when using Google Trends to assess “smell” queries and actual infection cases (r = 0.81) [18]. The interest in the above queries increased along with the number of new cases from 16/03/2020 to 11/05/2020 and from 27/08/2020 to 01/10/2020 (r = 0.93 and 0.87, respectively). From 02/04/2020 to 12/04/2020, the increase in queries outpaced the increase in actual COVID-19 cases by 2–5 days. The fact that the increase in the smell-related queries “outpaced” the increase in the new infection cases can imply that smell-impaired patients tend to study the problem through web search and only then opt for SARS-CoV-2 testing. We found that starting on 27/08/2020, the smell-related queries outnumbered the detected cases of infection as of 01/10/2020.

In this study, we analysed smell-related queries (hereafter “smell queries”) from Yandex Russian users in regions and large cities of Russia from 16/3/2020 to 21/2/2021 (49 weeks) and compared them with new cases of infection. In contrast to Google Trends, the Yandex.Wordstat service provides absolute rather than relative data, thus making it possible to compare queries with actual cases in absolute quantities, for example, coronavirus infection or drug consumption. We assume that smell-related queries can be markers of the spread of SARS-CoV-2 infection and can be used as a supplementary tool for evaluating the effectiveness of COVID-19 epidemic control measures.

## Materials and methods

### Study period

The study period lasted from 16/3/2020 to 21/2/2021 and was broken down into weeks.

### Databases

The data on new (confirmed) cases reported weekly in regions and large cities of were obtained using https://стопкоронавирус.рф (https://stopcoronavirus.rf) resources and Yandex and Johns Hopkins University services. The data (when required) were normalized on a scale from 0 to 100 by the maximum number of new (confirmed) cases per week during the study period.

### Databases of search queries

**Yandex.Wordstat** is the service providing information about queries made by Yandex users. For example, it helps determine the monthly number of people looking for a certain phrase and find queries similar in meaning to the entered phrase. The Yandex.Wordstat query statistics show how many times users entered their search-term-containing queries into the search box (the number of impressions). In our study, we estimated the number of users’ queries in Russia that included the word “smell”. We assume that this word could be used by users in key word combinations such as “loss of smell”, “smell loss recovery”, “treating smell loss”, “lost sense of smell”.

The data on the web-based smell query were received in absolute weekly values from 16/3/2020 – 22/3/2020 to 15/2/2021 – 21/2/2021.

## Results

### Dynamics of smell-related queries in the studied regions

Fig. 1 and Movie 1 (Supplementary movie 1) show changes in smell-related web queries from the 12^th^ week of 2020 (16/3/2020–22/3/2020) to the 7^th^ week of 2021 (15/2/2021–21/2/2021) (the total observation period consists of 49 weeks) in regions and large cities of Russia (85 regions). The data were first normalized to obtain the maximum number of queries per week over the entire period for each region. The maximum number of smell-related queries was recorded in most of the regions during the pandemic’s second wave in October-November, for example, in the Ivanovo, Kaluga, Moscow, and Ryazan regions from 2/11/2020-8/11/2020 and in Moscow from 23/11/2020-29/11/2020. The highest activity was observed during the first wave in several regions including Dagestan, Ingushetia, and North Ossetia from 27/4/2020 to 3/5/2020, Chechnya from 11/5/2020 to 17/5/2020, and Tyva from 22/6/2020 to 28/6/2020.

**Fig. 1.**
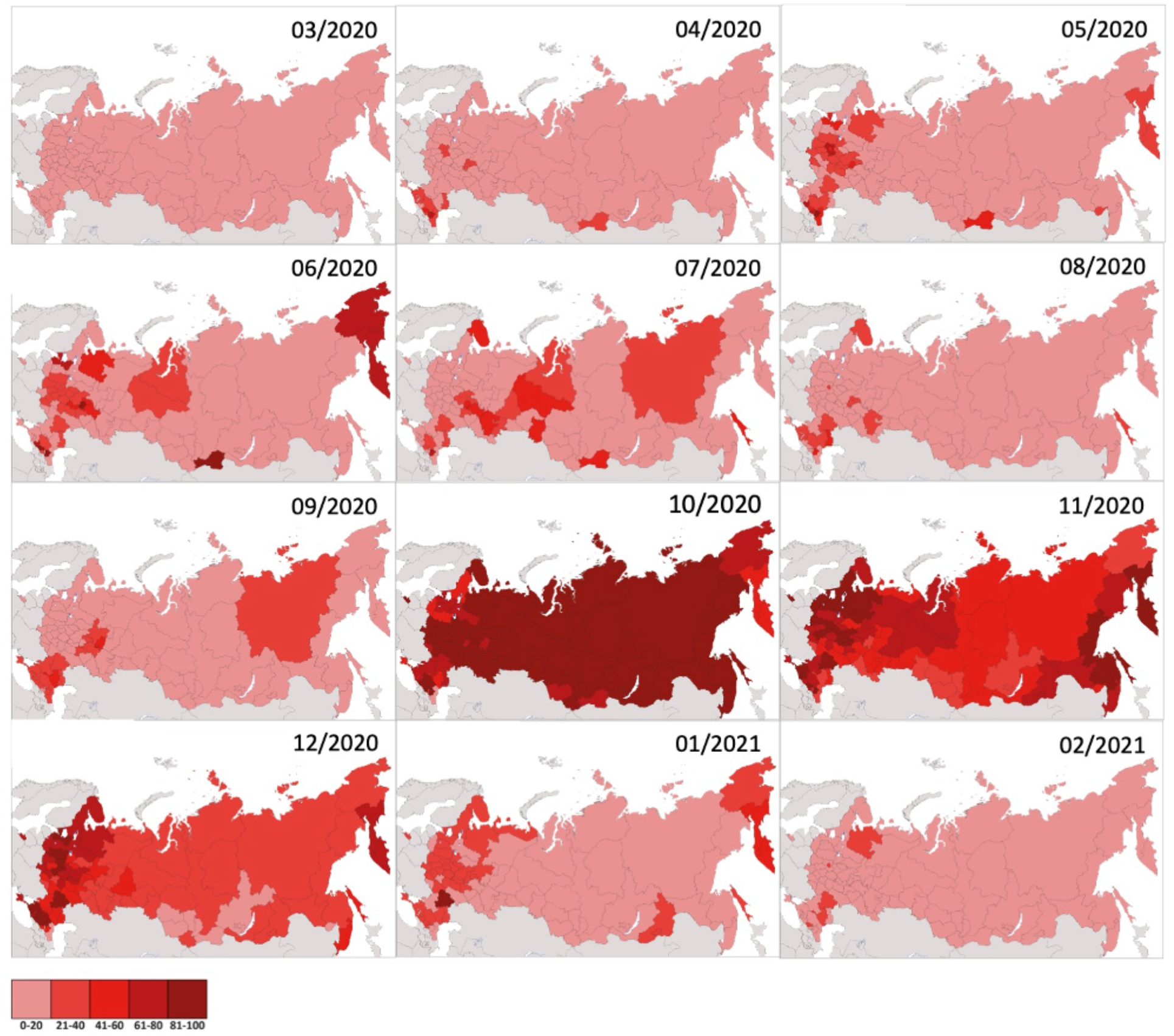
Heat map of changes in the number of smell-related queries when using the Yandex search engine from the 12^th^ week of 2020 (16/3/2020–22/3/2020) to the 7^th^ week of 2021 (15/2/2021– 21/2/2021) in Russia.

It was expected that the number of queries is strongly correlated (r>0.99) with the number of people living in the respective regions. When the smell-related queries (Supplementary Table 1) were normalized, the largest number of queries per population was found in Moscow (0.11 queries/person), St. Petersburg (0.10 queries/person), Nizhny Novgorod (0.09), Moscow, Novosibirsk, and Sverdlovsk regions (0.08). At the same time, Moscow, the Magadan Region, and the Altai Republic had the highest coronavirus infection incidence per person (>0.08). The smallest number of queries (0.01) was found in the Chechen Republic, the Republic of Ingushetia, and the Republic of Dagestan.

*The obtained data can imply specific behaviour patterns typical of populations in different regions of Russia during the coronavirus infection pandemic in terms of information search regarding one of the COVID-19 symptoms: loss of smell.*

### Correlations between the number of queries and new COVID-19 cases

We analysed correlations between the number of queries and new COVID-19 cases in regions and large cities of Russia (Supplementary Table 2, Fig. 2). The presented data show that Moscow is characterized by a very high correlation between queries and new cases of infection (r=0.96). The average (0.5<r<0.7) correlation between smell-related queries and new COVID-19 cases was detected for 25 regions (p<0.05). Thirty-three regions of Russia, including St.

**Fig. 2.**
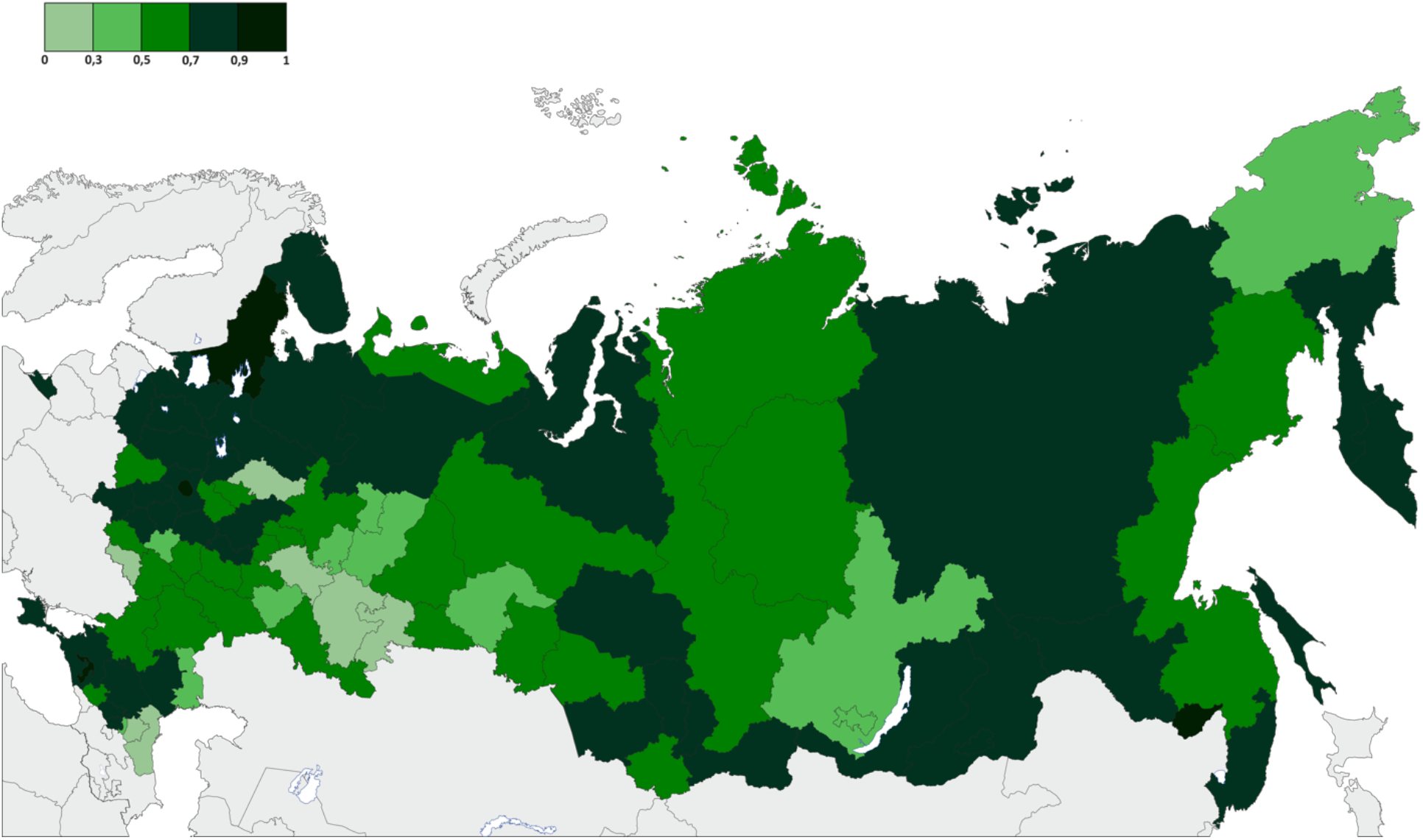
Correlations between the number of queries and new COVID-19 cases in Russia

Petersburg demonstrated a strong correlation (0.7<r<0.9) between the number of smell-related queries and new COVID-19 cases in the studied regions (p<0.05). For 26 regions, the correlation coefficient was less than 0.5 or p>0.05. Thus, in total, 70% of the regions and large cities of Russia (59 out of 85 regions) demonstrated a significantly strong correlation (r>0.7) between smell-related queries and confirmed COVID-19 cases; these data *suggest a strong relationship between the information search for one of the COVID-19 symptoms, loss of smell, and the confirmed cases of coronavirus infection.*

### The increase/decrease in the number of smell-related online queries precedes the increase/decrease in the number of new COVID-19 cases

We attempted to estimate the time gap between the online search of smell-related information by population and new cases of coronavirus infection in different regions of Russia. For this purpose, weekly queries and new cases of infection were normalized by the maximum number of queries and maximum number of new cases of infection per week over the entire observation period, respectively. For example, see Fig. 3; the number of smell-related queries and the number of new cases of infection before and after the data were normalized for Moscow and the Moscow and Vladimir Regions as a visual illustration of the obtained data. Then, we estimated the weekly increase in queries and new cases of infection by finding the difference between the number of queries (new cases of infection) per week and the number of queries (new cases of infection) during the previous week. The increase or decrease was considered significant if it was more than 2% and if the subsequent set of data had an upward or a downward trend. The analysis included only the regions that demonstrated a significant correlation (r>0.5) between smell-related queries and confirmed cases of infection.

**Fig. 3.**
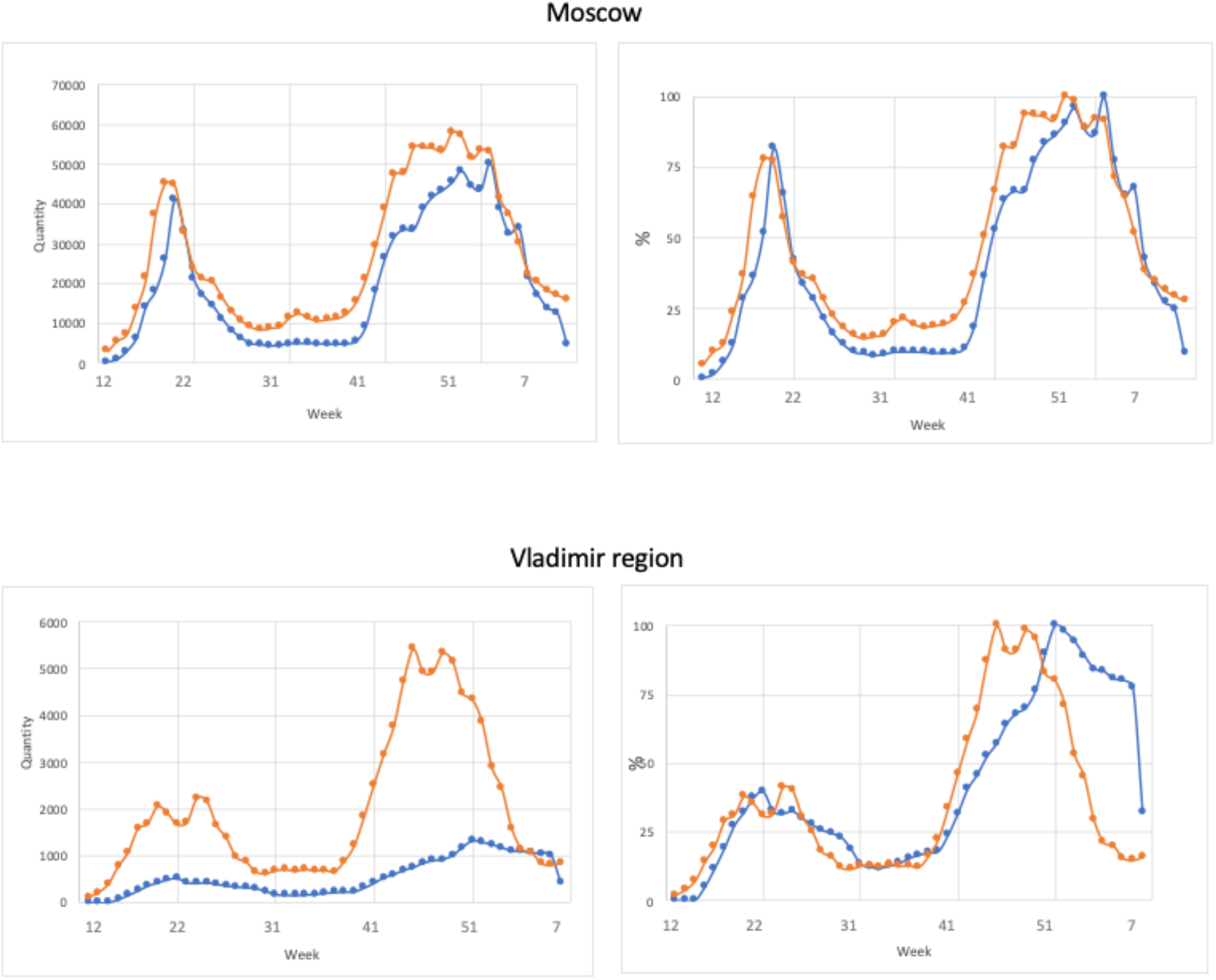
Dynamics in smell-related online queries and new cases of infection from the 12^th^ week of 2020 (16/3/2020–22/3/2020) to the 7^th^ week of 2021 (15/2/2021–21/2/2021) in Moscow and Vladimir region. On the left - the number of smell-related online queries and the number of new infection cases (absolute values); on the right – normalized values (%).

The results of the analysis are shown in Table 1 and Supplementary Table 3. The presented data show that during the first peak of coronavirus infection incidence in Russia, smell-related queries outpaced the increase in the number of new cases of infection by 1-2 weeks in 23 out of 59 regions of Russia. Such a relationship was not observed for the other regions.

**Table 1.** Weekly change in the increased number of online smell-related queries and the increased number of new cases of COVID-19

| Region of Russian Federation | 13-22 weeks | 34-40 weeks |
| --- | --- | --- |
| Altai Territory |  | 1 |
| Amursk Region |  | 1 |
| Arkhangelsk Region | 1 | 1 |
| Bryansk Region |  |  |
| Vladimir Region | 2 | 2 |
| Volgograd Region | 2 | 3 |
| Voronezh Region | 1 | 3 |
| Ivanovo Region |  | 2 |
| Kaliningrad Region |  | 1 |
| Kaluga Region | 2 |  |
| Karachay-Cherkess Republic | 2 |  |
| Kemerovo Territory |  | 2 |
| Kirov Region |  | 3 |
| Kostroma Region |  | 3 |
| Krasnodar Territory | 1 | 3 |
| Krasnoyarsk Territory | 1 | 2 |
| Republic of Crimea |  |  |
| Kurgan Region |  | 1 |
| Kursk Region | 5 | 1 |
| Leningrad Region | 1 | 1 |
| Magadan Region |  | 1 |
| Moscow | 1 | 2 |
| Moscow Region | 5 | 4 |
| Murmansk Region |  | 2 |
| Nenets Autonomous District | 2 | 2 |
| Nizhny Novgorod Region |  | 3 |
| Novgorod Region | 1 | 2 |
| Novosibirsk Region | 1 | 2 |
| Omsk Region |  | 1 |
| Orenburg Region |  | 2 |
| Oryol Region | 1 | 1 |
| Penza Region |  | 4 |
| Primorsky Territory |  | 4 |
| Republic of Altai |  | 1 |
| Republic of Kalmykia |  |  |
| Republic of Karelia |  | 2 |
| Mari El Republic | 1 | 1 |
| Republic of Mordovia |  | 1 |
| Republic of Sakha (Yakutia) | 2 | 2 |
| Republic of North Ossetia-Alania | 2 | 6 |
| Republic of Tyva | 5 | 2 |
| Rostov Region |  | 1 |
| Ryazan Region |  | 2 |
| St. Petersburg | 2 | 4 |
| Saratov Region |  | 1 |
| Sakhalin Region |  | 3 |
| Sverdlovsk Region |  | 5 |
| Sevastopol |  |  |
| Smolensk Region | 1 | 2 |
| Stavropol Territory | 2 |  |
| Tambov Region |  | 1 |
| Tver Region | 8 | 8 |
| Tomsk Region |  | 2 |
| Tula Region | 1 | 1 |
| Ulyanovsk Region |  | 2 |
| Khanty-Mansi Autonomous Area |  | 1 |
| Chuvash Republic | 1 |  |
| Yamal-Nenets Autonomous District |  | 1 |
| Yaroslavl Region | 1 |  |

During the second peak of coronavirus infection incidence in Russia, the increase in the number of smell-related queries outpaced the increase in the number of new cases of infection by 1-2 weeks in 36 regions of Russia, including Moscow. In 14 regions, queries outnumbered new cases of infection, and they were ahead by more than 2 weeks. Two regions of Russia did not demonstrate such patterns during the first and second waves.

### The relationship between queries and new infection cases as a supplementary tool for evaluation of the effectiveness of COVID-19 epidemic control measures

Since the Yandex.Wordstat provides queries in absolute rather than relative numbers, we estimated the relative increase in the number of queries during the second peak of coronavirus infection incidence as the effectiveness indicator for COVID-19 epidemic control measures in regions of Russia. For this purpose, we compared the relationships between smell-related queries and new COVID-19 cases between two periods: when the number of new infections in the studied regions was minimal for several weeks (the plateau period from 6/7/2020 (28^th^ week) – 12/7/2020 to 14/9/2020 – 20/9/2020 (38^th^ week)) and when the number of infections had an upward trend, rising to the maximum number (the peak period from 21/9/2020-27/9/2020 (39^th^ week) to 30/11/2020 – 6/12/2020 (49^th^ week)).

Fig. 4 and Table 2 show the results of the calculation of the query/new infection case ratio for 45 regions between the peak period and the plateau period. The obtained data show that, except for Moscow, St. Petersburg, and the Tver Region, the ratio of queries to the number of new cases of infection increased during the second peak of coronavirus infection incidence compared to the plateau period; in 24 regions, the increase in queries was more than 100%.

**Fig. 4.**
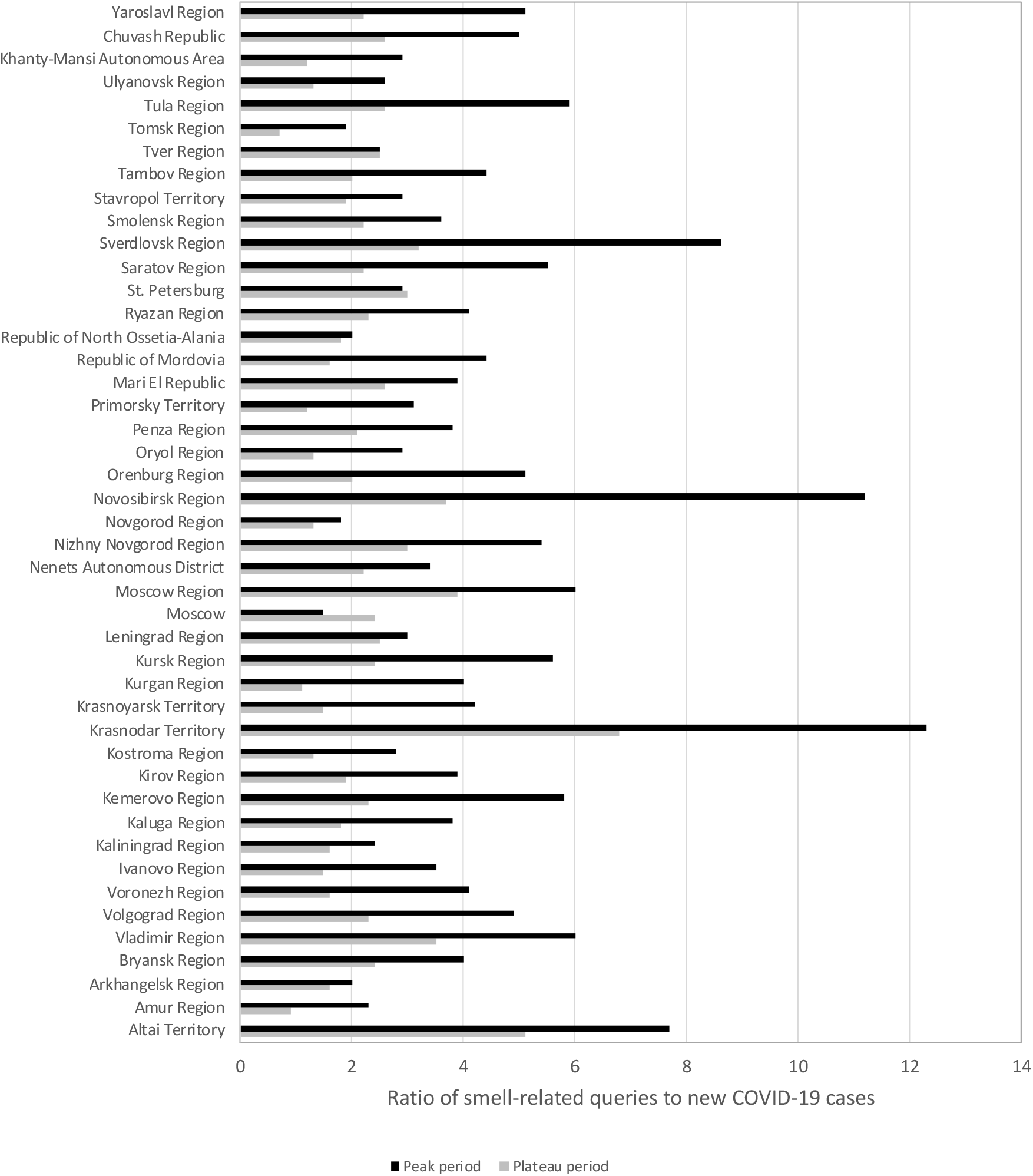
Comparison of the ratio of smell-related queries to new COVID-19 cases in different periods. The plateau period — the number of new cases of infection in the studied regions was minimal during several weeks: from 6/7/2020–12/7/2020 (28^th^ week) to 14/9/2020–20/9/2020 (38^th^ week). The peak period — the number of new cases of infection was characterized by an upward trend, rising to a maximum: from 21/9/2020–27/9/2020 (39^th^ week) to 30/11/2020–6/12/2020 (49^th^ week)).

**Table 2.** The ratio of smell-related queries to new cases of COVID-19 between two periods of plateau and peak

| Region of Russian Federation | Queries/cases (total) | Queries/cases (plateau) | Queries/cases (peak) | Peak/plateau |
| --- | --- | --- | --- | --- |
| Moscow | 1.7±0.6 | 2.4±0.3 | 1.5±0.3 | 63% |
| St. Petersburg | 3±1.8 | 3±0.8 | 2.9±1.4 | 97% |
| Tver Region | 2.9±1.8 | 2.5±0.4 | 2.5±0.3 | 100% |
| Republic of North Ossetia-Alania | 1.6±0.8 | 1.8±0.9 | 2±0.7 | 111% |
| Leningrad Region | 3.2±1.8 | 2.5±0.5 | 3±0.5 | 120% |
| Arkhangelsk Region | 1.8±0.7 | 1.6±0.4 | 2±0.4 | 125% |
| Novgorod Region | 1.4±0.7 | 1.3±0.4 | 1.8±0.3 | 138% |
| Kaliningrad Region | 1.6±1 | 1.6±0.3 | 2.4±0.8 | 150% |
| Mari El Republic | 2.4±1.1 | 2.6±0.4 | 3.9±0.8 | 150% |
| Altai Territory | 16.8±55.9 | 5.1±1.3 | 7.7±4 | 151% |
| Stavropol Territory | 2.6±1.6 | 1.9±0.3 | 2.9±0.6 | 153% |
| Moscow Region | 3.6±1.8 | 3.9±0.8 | 6±1.4 | 154% |
| Nenets Autonomous District | 3.4±5.4 | 2.2±3.4 | 3.4±1.2 | 155% |
| Smolensk Region | 2.4±1.2 | 2.2±0.9 | 3.6±0.5 | 164% |
| Bryansk Region | 2.5±1.2 | 2.4±0.7 | 4±0.6 | 167% |
| Vladimir Region | 4±1.7 | 3.5±0.8 | 6±0.6 | 171% |
| Ryazan Region | 2.9±1.2 | 2.3±0.5 | 4.1±0.6 | 178% |
| Nizhny Novgorod Region | 3.2±1.5 | 3±0.6 | 5.4±0.7 | 180% |
| Krasnodar Territory | 7.5±4.1 | 6.8±1.5 | 12.3±1.9 | 181% |
| Penza Region | 2.7±1.6 | 2.1±0.4 | 3.8±1 | 181% |
| Chuvash Republic | 3.2±1.6 | 2.6±0.5 | 5±1.3 | 192% |
| Ulyanovsk Region | 1.8±1.2 | 1.3±0.3 | 2.6±0.7 | 200% |
| Kirov Region | 2.2±1.5 | 1.9±0.7 | 3.9±1.9 | 205% |
| Kaluga Region | 2.3±1.1 | 1.8±0.5 | 3.8±0.9 | 211% |
| Volgograd Region | 3±2.9 | 2.3±0.6 | 4.9±1.9 | 213% |
| Kostroma Region | 2.0±1.3 | 1.3±0.6 | 2.8±0.8 | 215% |
| Tambov Region | 2.5±1.4 | 2±0.5 | 4.4±1.3 | 220% |
| Oryol Region | 1.7±0.9 | 1.3±0.3 | 2.9±0.4 | 223% |
| Tula Region | 3.3±1.7 | 2.6±0.8 | 5.9±0.9 | 227% |
| Yaroslavl Region | 2.9±1.6 | 2.2±0.5 | 5.1±1.1 | 232% |
| Ivanovo Region | 2±1.3 | 1.5±0.4 | 3.5±0.8 | 233% |
| Kursk Region | 10.3±15.5 | 2.4±1 | 5.6±1.6 | 233% |
| Khanty-Mansi Autonomous Area | 2.1±1.6 | 1.2±0.4 | 2.9±1.1 | 242% |
| Saratov Region | 3.2±2.8 | 2.2±0.4 | 5.5±2.1 | 250% |
| Kemerovo Territory | 6.4±11.1 | 2.3±0.6 | 5.8±2.8 | 252% |
| Orenburg Region | 2.7±2.1 | 2±0.8 | 5.1±2.5 | 255% |
| Amursk Region | 1.9±2.6 | 0.9±0.3 | 2.3±0.6 | 256% |
| Voronezh Region | 2.7±2.2 | 1.6±0.6 | 4.1±1.8 | 256% |
| Primorsky Territory | 1.9±1.7 | 1.2±0.4 | 3.1±0.7 | 258% |
| Sverdlovsk Region | 5.4±6.6 | 3.2±0.9 | 8.6±2 | 269% |
| Tomsk Region | 1.6±2.1 | 0.7±0.2 | 1.9±0.6 | 271% |
| Republic of Mordovia | 2.8±1.4 | 1.6±0.3 | 4.4±1.2 | 275% |
| Krasnoyarsk Territory | 2.5±1.7 | 1.5±0.6 | 4.2±1.5 | 280% |
| Novosibirsk Region | 6.2±4.9 | 3.7±1.3 | 11.2±4.7 | 303% |
| Kurgan Region | 3.2±5.1 | 1.1±0.4 | 4±1.8 | 364% |

Note. The plateau period - the number of new cases of infection in the studied regions was the smallest during several weeks: from 6/7/2020–12/7/2020 (28<sup>th</sup> week) to 14/9/2020–20/9/2020 (38<sup>th</sup> week). The peak period – the number of new cases of infection had an upward trend, rising to the maximum number: from 21/9/2020–27/9/2020) (39<sup>th</sup> week) to 30/11/2020–6/12/2020 (49<sup>th</sup> week).

Furthermore, in these regions, we assessed the popularity of queries related to levofloxacin, an antibiotic which was mentioned in the guidelines of the Ministry of Health of Russia as an agent administered to treat bacterial infections in COVID-19 patients (Fig. 5 and Supplementary Table 4). As seen from the data, we can point out two groups where the ratio of levofloxacin-related queries to the total number of new cases of infection was less than one (Group 1) or larger than one (Group 2). The first group includes Moscow and St. Petersburg, where the ratio of queries to the number of new cases of infection decreased during the peak period compared to the plateau period.

**Fig. 5.**
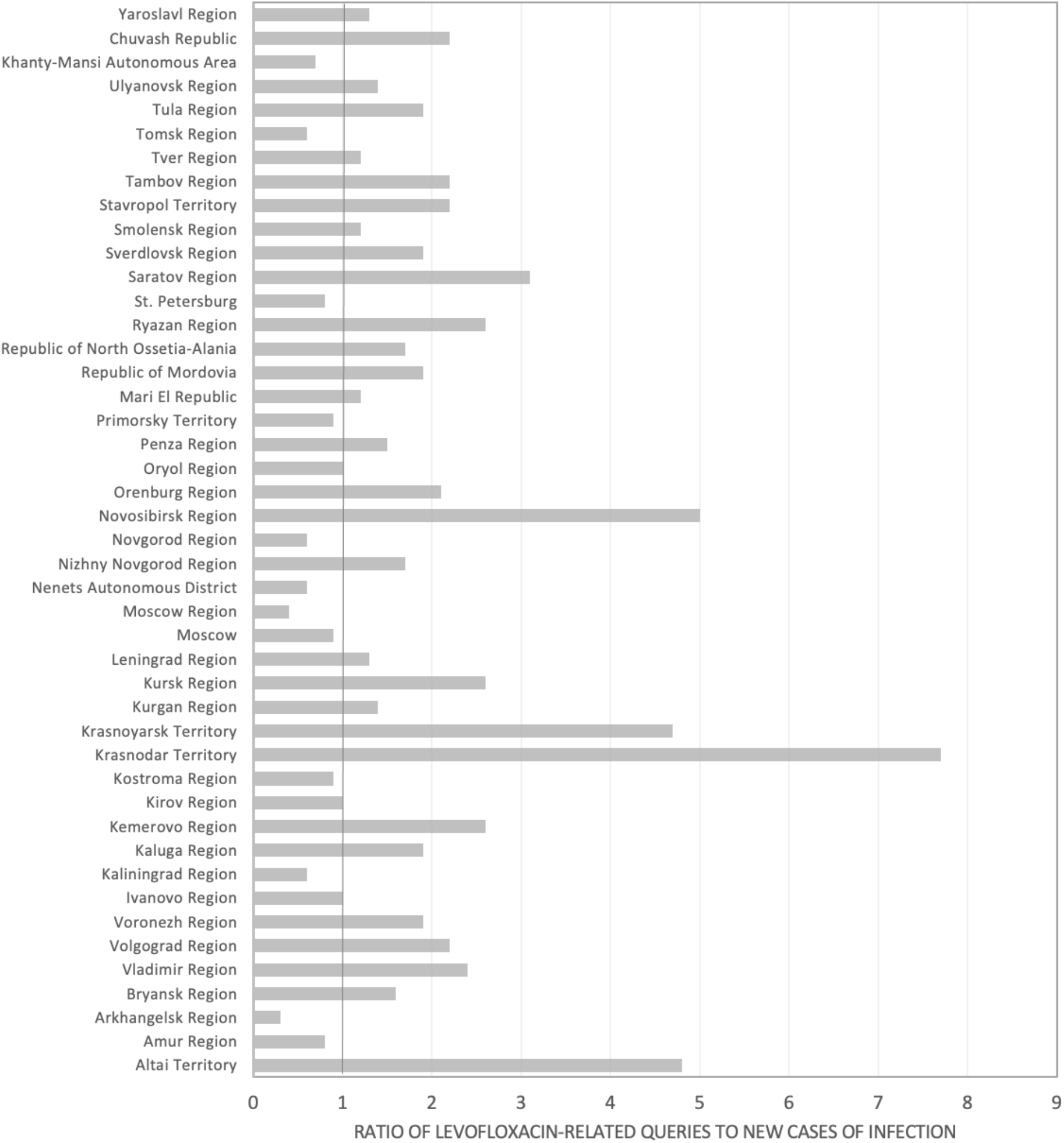
The ratio of levofloxacin-related queries to new cases of infection in 45 regions of Russia.

No significant relationship was found between the increase in smell-related queries during the peak period with the cut-off point set at 100% and the increase in levofloxacin-related queries compared to the number of new cases of infection (p=0.1690). However, when the cut-off point was set at 160%, a significant relationship (p=0.0216) was found between the increase in smell-related queries during the peak period and the increase in levofloxacin-related search activity. In other words, in the regions where the increase in queries compared to the number of new cases of infection during the second peak of coronavirus infection incidence increased by more than 160%, the levofloxacin-related search activity was significantly higher than in regions where the number of queries increased by less than 160% compared to the number of new cases of infection.

## Discussion

This study demonstrated a significant strong correlation (r>0.7) between the number of smell-related queries in Yandex and new COVID-19 cases in 59 regions and 85 large cities of Russia (70%). The obtained results are consistent with our previous data that revealed a strong correlation between smell queries and new infection cases (r = 0.81) in Russia using Google Trends [18]. Higgins et al. [22] also pointed out that worldwide search queries related to shortness of breath, anosmia, dysgeusia and ageusia, headache, chest pain, and sneezing had a strong correlation (r>0.60; p<0.001) both with daily new confirmed cases and with the number of deaths caused by COVID-19. Similar results were obtained by Walker et al [23], who found a strong correlation between daily RSV (relative search volume) associated with loss of smell, the daily increase in COVID-19 cases, and deaths, ranging from 0.633 to 0.952 (p<0.05) in several countries.

In addition, the obtained data show that during the first peak of the coronavirus infection incidence in Russia, the increase in smell-related queries outpaced the increase in new infection cases by 1-2 weeks in 23 out of 59 regions. During the second peak of the coronavirus infection incidence in Russia, the increase in the number of smell-related queries outpaced the increase in the number of new infection cases by 1-2 weeks in 36 regions of Russia, including Moscow. In 14 regions, the increase in queries outpaced the increase in new infection cases by more than 2 weeks. The time interval between the onset of COVID-19-associated symptoms and their actual detection was also shown in a previous study [24].

An important question raised in our study is whether the smell-related queries are primarily attributed to the queries from users with COVID-19 or they can be explained by other time-related reasons such as seasonal diseases, allergies, or infodemic (a rapid spread of information) when users who do not experience COVID-19 symptoms, including loss of smell, try to find more information about the disease [24].

When analysing the Yandex.Wordstat data, we did not observe such peaks in queries related to loss of smell during the respective period in previous years. The assumption that symptom-free occasional users are interested in COVID-19 symptoms can be challenged by several arguments. For example, a comparative study conducted in Israel [25] showed that patients suspected of having COVID-19 and having positive COVID-19 results (68%) demonstrated changes in smell almost ten times as frequently as patients with negative COVID-19 (8%) results. Our study did not detect any increase in queries compared to new cases of infection during the second wave of infection in Moscow, St. Petersburg, and the Tver Region, the total population of which accounts for 50% of the population of 24 regions; there was a 100-250% increase in the number of queries compared to the number of new cases of infection. In addition, for Moscow, the ratio of queries to new cases of infection decreased from 2.4±0.3 to 1.5±0.3, while for St. Petersburg and the Tver Region, the ratio of queries to new cases of infection remained unchanged. Moreover, from 16/11/2020 – 22/11/2020 to 25/1/2021 – 31/1/2021, in Moscow, the number of queries exceeded the number of new cases of infection by only 10-20% (Fig. 3). Therefore, it is quite probable that smell-related queries are generated by people suffering from loss of smell, which is primarily associated with COVID-19.

The existing difference (100-250%) between the number of queries and confirmed COVID-19 cases during the second peak of coronavirus infection incidence can be explained by the fact that some of the users who suffered from loss of smell and who searched the Internet for information did not see a doctor in healthcare facilities. Apparently, in this case, patients can opt for self-treatment and look for information about methods of treatment of coronavirus infection. One of these antibiotics is levofloxacin, which was mentioned in the guidelines of the Ministry of Health of Russia as an agent to treat bacterial infections in COVID-19 patients. A shortage of this antibiotic was reported in Russia starting in November 2021. We found that in regions where the queries increased by 160% compared to the number of new cases of infection during the second wave, the levofloxacin-related search activity was also significantly higher than in regions where the number of queries increased by less than 160% compared to the number of new cases of infection.

Our study shows that the analysis of search queries in Yandex.Wordstat confirms the timewise relationship: internet users first look for information about their initial COVID-19 symptoms (loss of smell) and then have their disease confirmed. The presented data demonstrate that the increase (decrease) in the number of smell-related queries precedes the increase (decrease) in the number of infections by several weeks. Therefore, the ratio of queries to new cases of infection can be used to estimate the actual number of patients with recent coronavirus infection. For example, from 16/11/2020 – 22/11/2020 to 25/1/2021 – 31/1/2021, the queries in Moscow outnumbered the new infections by only 10-20%. This suggests an effective policy targeted at COVID-19 epidemic control measures in Moscow, when all the people affected by COVID-19 were detected in a timely manner.

We assume that the increase in sudden interest in smell changes can be used as a valuable, minimally invasive indicator of coronavirus spread among populations as well as a tool for evaluating the effectiveness of COVID-19 epidemic control measures.

## Data Availability

All data generated or analysed during this study are included in this published article (and its supplementary information files).

## Supplementary Information

Supplementary Table 1. Statistics of smell-related queries in Russia

Supplementary Table 2. The correlation between the number of queries and new cases of COVID-19 in Russia.

Supplementary Table 3. Weekly change in the increased number of online smell-related queries and the increased number of new cases of COVID-19.

Supplementary Table 4. Statistics of levofloxacin-related queries in Russia.

## Notes

### Competing Interest Statement

The authors have declared no competing interest.

### Funding Statement

No funding provided for this study.

### Author Declarations

ll relevant ethical guidelines from Central Research Institute of Epidemiology of the Federal Service for Customers Rights Protection and Human Well-Being Surveillance have been followed. All necessary patient/participant consent has been obtained and the appropriate institutional forms have been archived.

