## Supplementary table 1 for "Online queries as a criterion for evaluation of the epidemiological status and effectiveness of COVID-19 epidemic control measures"

**Supplementary Table 1. Statistics of smell-related queries in Russia**

| Region of the Russian Federation | Population. people | Total (new cases) | Maximum per week (new cases) | Total (queries) | Maximum per week (queries) | Queries/people |
|---|---|---|---|---|---|---|
| Altai Territory | 2 296 773 | 15954 | 1252 | 98146 | 11064 | 0.04 |
| Amur Region | 781 850 | 20511 | 1165 | 29495 | 2562 | 0.04 |
| Arkhangelsk Region | 1 128 099 | 34158 | 1593 | 53708 | 3032 | 0.05 |
| Astrakhan Region | 997 685 | 27008 | 1297 | 38105 | 1954 | 0.04 |
| Belgorod Region | 1 543 087 | 30201 | 1304 | 81706 | 7359 | 0.05 |
| Bryansk Region | 1 183 228 | 31714 | 1411 | 76022 | 5101 | 0.06 |
| Vladimir Region | 1 343 194 | 25969 | 1307 | 96504 | 5417 | 0.07 |
| Volgograd Region | 2 476 100 | 47180 | 1927 | 123163 | 9252 | 0.05 |
| Vologda Region | 1 151 751 | 47180 | 1927 | 62594 | 4790 | 0.05 |
| Voronezh Region | 2 307 100 | 64411 | 2736 | 143372 | 11045 | 0.06 |
| Jewish Autonomous Region | 156 492 | 4281 | 322 | 4419 | 387 | 0.03 |
| Trans-Baikal Territory | 1 054 071 | 37341 | 1876 | 39475 | 3345 | 0.04 |
| Ivanovo Region | 987 694 | 29393 | 1317 | 53471 | 3798 | 0.05 |
| Irkutsk Region | 2 375 640 | 53018 | 1933 | 116682 | 13497 | 0.05 |
| Kabardino-Balkarian Republic | 870 197 | 20178 | 713 | 21299 | 885 | 0.02 |
| Kaliningrad Region | 1 019 601 | 26900 | 1523 | 38943 | 2918 | 0.04 |
| Kaluga Region | 1 000 604 | 28473 | 1199 | 68360 | 4446 | 0.07 |
| Kamchatka Territory | 311 985 | 13106 | 571 | 12779 | 710 | 0.04 |
| Karachay-Cherkess Republic | 465 592 | 18568 | 675 | 7982 | 392 | 0.02 |
| Kemerovo Region | 2 634 376 | 30891 | 1377 | 122241 | 12347 | 0.05 |
| Kirov Region | 1 251 182 | 35041 | 1753 | 71534 | 4285 | 0.06 |
| Kostroma Region | 628 972 | 1202 | 100 | 35937 | 3011 | 0.06 |
| Krasnodar Territory | 5 689 538 | 36679 | 1378 | 287423 | 16303 | 0.05 |
| Krasnoyarsk Territory | 2 857 567 | 60689 | 2354 | 151088 | 13673 | 0.05 |
| Republic of Crimea | 1 903 707 | 34772 | 2466 | 94285 | 6413 | 0.05 |
| Kurgan Region | 819 299 | 16787 | 758 | 33160 | 3109 | 0.04 |

| | | | | | | |
|---|---|---|---|---|---|---|
| Kursk Region | 1 098 361 | 16787 | 758 | 68311 | 4685 | 0.06 |
| Leningrad Region | 1 894 038 | 34158 | 1593 | 96979 | 5217 | 0.05 |
| Lipetsk Region | 1 128 181 | 23352 | 1168 | 72541 | 5308 | 0.06 |
| Magadan Region | 138 991 | 13106 | 571 | 4488 | 338 | 0.03 |
| Moscow | 12 636 312 | 960652 | 50304 | 1326923 | 58136 | 0.11 |
| Moscow Region | 7 713 326 | 206936 | 10919 | 638954 | 30305 | 0.08 |
| Murmansk Region | 733 158 | 45650 | 2006 | 40040 | 2212 | 0.05 |
| Nenets Autonomous District | 44 404 | 989 | 63 | 1953 | 213 | 0.04 |
| Nizhny Novgorod Region | 3 177 816 | 92136 | 3509 | 295507 | 15834 | 0.09 |
| Novgorod Region | 593 232 | 26185 | 1472 | 32214 | 2378 | 0.05 |
| Novosibirsk Region | 2 786 412 | 34437 | 1262 | 214460 | 20406 | 0.08 |
| Omsk Region | 1 904 294 | 39799 | 1776 | 101265 | 7793 | 0.05 |
| Orenburg Region | 1 945 307 | 36509 | 1328 | 100097 | 8077 | 0.05 |
| Oryol Region | 725 725 | 29213 | 1260 | 52590 | 3213 | 0.07 |
| Penza Region | 1 292 211 | 36276 | 1598 | 86159 | 5127 | 0.07 |
| Perm Territory | 2 580 639 | 44389 | 2323 | 135767 | 9914 | 0.05 |
| Primorsky Territory | 1 879 486 | 39248 | 1682 | 74032 | 5671 | 0.04 |
| Pskov Region | 621 028 | 30923 | 2889 | 31816 | 2789 | 0.05 |
| Republic of Adygea | 463 091 | 13615 | 702 | 12929 | 958 | 0.03 |
| Republic of Altai | 221 050 | 41949 | 1649 | 7086 | 690 | 0.03 |
| Republic of Bashkortostan | 4 016 481 | 26994 | 1177 | 202409 | 12978 | 0.05 |
| Republic of Buryatia | 986 132 | 32498 | 1902 | 31214 | 3475 | 0.03 |
| Republic of Dagestan | 3 132 368 | 18568 | 675 | 42914 | 4646 | 0.01 |
| Republic of Ingushetia | 515 970 | 14517 | 620 | 6036 | 618 | 0.01 |
| Republic of Kalmykia | 270 139 | 18509 | 747 | 8201 | 496 | 0.03 |
| Republic of Karelia | 609 439 | 39217 | 3176 | 30348 | 2628 | 0.05 |
| Komi Republic | 813 859 | 38541 | 2123 | 40381 | 2982 | 0.05 |
| Mari El Republic | 676 184 | 11385 | 373 | 27274 | 1432 | 0.04 |
| Republic of Mordovia | 779 753 | 17207 | 627 | 52523 | 3078 | 0.07 |
| Republic of Sakha (Yakutia) | 984 703 | 32017 | 1559 | 23475 | 1550 | 0.02 |

| | | | | | | |
|---|---|---|---|---|---|---|
| Republic of North Ossetia-Alania | 693 449 | 15322 | 642 | 22308 | 1284 | 0.03 |
| Republic of Tatarstan | 3 894 507 | 17367 | 716 | 215640 | 12905 | 0.06 |
| Republic of Tyva | 330 327 | 15596 | 950 | 6448 | 360 | 0.02 |
| Republic of Khakassia | 532 266 | 21060 | 1267 | 23472 | 2345 | 0.04 |
| Rostov Region | 4 185 233 | 69683 | 2750 | 209825 | 15327 | 0.05 |
| Ryazan Region | 1 098 998 | 29213 | 1260 | 80771 | 4832 | 0.07 |
| Samara Region | 3 155 390 | 45398 | 2145 | 220062 | 15226 | 0.07 |
| St. Petersburg | 5 388 759 | 356376 | 26333 | 553274 | 28169 | 0.10 |
| Saratov Region | 2 396 961 | 47197 | 1892 | 134832 | 10076 | 0.06 |
| Sakhalin Region | 485 627 | 20959 | 1028 | 16605 | 994 | 0.03 |
| Sverdlovsk Region | 4 291 886 | 75487 | 2834 | 347030 | 23788 | 0.08 |
| Sevastopol | 513 149 | 11215 | 698 | 20795 | 1899 | 0.04 |
| Smolensk Region | 921 620 | 25150 | 1300 | 52606 | 3183 | 0.06 |
| Stavropol Territory | 2 794 625 | 46038 | 1713 | 110376 | 5412 | 0.04 |
| Tambov Region | 995 761 | 24904 | 1123 | 57875 | 3683 | 0.06 |
| Tver Region | 1 246 759 | 32079 | 1652 | 75080 | 4644 | 0.06 |
| Tomsk Region | 1 070 544 | 29619 | 1472 | 41029 | 3957 | 0.04 |
| Tula Region | 1 450 675 | 31603 | 1452 | 100600 | 5808 | 0.07 |
| Tyumen Region | 3 778 024 | 75487 | 2834 | 77417 | 6059 | 0.02 |
| Republic of Udmurtia | 1 493 700 | 27195 | 1530 | 68232 | 5123 | 0.05 |
| Ulyanovsk Region | 1 219 240 | 46401 | 2014 | 72381 | 4153 | 0.06 |
| Khabarovsk Territory | 1 302 918 | 47876 | 2394 | 53758 | 3904 | 0.04 |
| Khanty-Mansi Autonomous Area | 1 688 378 | 50091 | 2116 | 92091 | 6514 | 0.05 |
| Chelyabinsk Region | 3 444 025 | 48371 | 2176 | 200202 | 16099 | 0.06 |
| Chechen Republic | 1 500 490 | 11167 | 995 | 9972 | 379 | 0.01 |
| Chuvash Republic | 1 208 806 | 20787 | 902 | 62011 | 3077 | 0.05 |
| Chukotka Autonomous District | 49 300 | 672 | 52 | 1174 | 80 | 0.02 |
| Yamal-Nenets Autonomous District | 547 111 | 36620 | 1399 | 29068 | 2126 | 0.05 |
| Yaroslavl Region | 1 242 356 | 31111 | 1334 | 88558 | 6221 | 0.07 |
