## Supplementary table 2 for "Online queries as a criterion for evaluation of the epidemiological status and effectiveness of COVID-19 epidemic control measures"

**Supplementary Table 2. The correlation between the number of queries and new cases of COVID-19 in Russia**

| Region of Russian Federation | Correlations | p-value | Region | Correlations | p-value | Region | Correlations | p-value |
|---|---|---|---|---|---|---|---|---|
| Altai Territory | 0.8 | 0.0000 | Magadan Region | 0.5 | 0.0000 | Republic of Tyva | 0.8 | 0.0000 |
| Amur Region | 0.8 | 0.0103 | Moscow | 1.0 | 0.0000 | Republic of Khakassia | 0.7 | 0.4478 |
| Arkhangelsk Region | 0.7 | 0.0000 | Moscow Region | 0.7 | 0.0000 | Rostov Region | 0.6 | 0.0000 |
| Astrakhan Region | 0.3 | 0.0019 | Murmansk Region | 0.8 | 0.0256 | Ryazan Region | 0.7 | 0.0000 |
| Belgorod Region | 0.3 | 0.0000 | Nenets Autonomous District | 0.6 | 0.0011 | Samara Region | 0.3 | 0.0000 |
| Bryansk Region | 0.7 | 0.0000 | Nizhny Novgorod Region | 0.7 | 0.0000 | St. Petersburg | 0.7 | 0.0000 |
| Vladimir Region | 0.6 | 0.0000 | Novgorod Region | 0.8 | 0.0242 | Saratov Region | 0.5 | 0.0000 |
| Volgograd Region | 0.5 | 0.0000 | Novosibirsk Region | 0.6 | 0.0000 | Sakhalin Region | 0.8 | 0.0064 |
| Vologda Region | 0.7 | 0.0530 | Omsk Region | 0.5 | 0.0000 | Sverdlovsk Region | 0.6 | 0.0000 |
| Voronezh Region | 0.5 | 0.0000 | Orenburg Region | 0.5 | 0.0000 | Sevastopol | 0.7 | 0.0005 |
| Jewish Autonomous Region | 0.9 | 0.6978 | Oryol Region | 0.7 | 0.0001 | Smolensk Region | 0.6 | 0.0000 |
| Trans-Baikal Territory | 0.8 | 0.5744 | Penza Region | 0.6 | 0.0000 | Stavropol Territory | 0.8 | 0.0000 |
| Ivanovo Region | 0.5 | 0.0002 | Perm Territory | 0.3 | 0.0000 | Tambov Region | 0.5 | 0.0000 |
| Irkutsk Region | 0.4 | 0.0035 | Primorsky Territory | 0.7 | 0.0003 | Tver Region | 0.8 | 0.0000 |
| Kabardino-Balkarian Republic | 0.7 | 0.3613 | Pskov Region | 0.8 | 0.7701 | Tomsk Region | 0.8 | 0.0211 |
| Kaliningrad Region | 0.7 | 0.0092 | Republic of Adygea | 0.9 | 0.4181 | Tula Region | 0.7 | 0.0000 |
| Kaluga Region | 0.7 | 0.0000 | Republic of Altai | 0.6 | 0.0000 | Tyumen Region | 0.3 | 0.8636 |
| Kamchatka Territory | 0.8 | 0.6483 | Republic of Bashkortostan | 0.2 | 0.0000 | Republic of Udmurtia | 0.3 | 0.0000 |
| Karachay-Cherkess Republic | 0.6 | 0.0000 | Republic of Buryatia | 0.7 | 0.7604 | Ulyanovsk Region | 0.6 | 0.0004 |
| Kemerovo Region | 0.7 | 0.0000 | Republic of Dagestan | -0.2 | 0.0010 | Khabarovsk Territory | 0.6 | 0.3335 |

| | | | | | | | | |
|---|---|---|---|---|---|---|---|---|
| Kirov Region | 0.6 | 0.0000 | Republic of Ingushetia | 0.3 | 0.0000 | Khanty-Mansi Autonomous Area | 0.6 | 0.0000 |
| Kostroma Region | 0.8 | 0.0000 | Republic of Kalmykia | 0.8 | 0.0000 | Chelyabinsk Region | 0.1 | 0.0000 |
| Krasnodar Territory | 0.7 | 0.0000 | Republic of Karelia | 0.9 | 0.0160 | Chechen Republic | 0.2 | 0.5552 |
| Krasnoyarsk Territory | 0.6 | 0.0001 | Komi Republic | 0.8 | 0.6031 | Chuvash Republic | 0.5 | 0.0000 |
| Republic of Crimea | 0.8 | 0.0000 | Mari El Republic | 0.6 | 0.0000 | Chukotka Autonomous District | 0.4 | 0.0005 |
| Kurgan Region | 0.5 | 0.0015 | Republic of Mordovia | 0.7 | 0.0000 | Yamal-Nenets Autonomous District | 0.7 | 0.0071 |
| Kursk Region | 0.6 | 0.0000 | Republic of Sakha (Yakutia) | 0.7 | 0.0010 | Yaroslavl Region | 0.7 | 0.0000 |
| Leningrad Region | 0.7 | 0.0000 | Republic of North Ossetia-Alania | 0.7 | 0.0002 | | | |
| Lipetsk Region | 0.3 | 0.0000 | Republic of Tatarstan | 0.1 | 0.0000 | | | |
