## Supplementary table 3 for "Online queries as a criterion for evaluation of the epidemiological status and effectiveness of COVID-19 epidemic control measures"

**Supplementary Table 3. Weekly change in the increased number of online smell-related queries and the increased number of new cases of COVID-19**

| Region of Russian Federation | The beginning of the increase in the number of queries (week) | The beginning of the increase in the number of new cases of infection (week) | Change (weekly) | The beginning of the increase in the number of queries (week) | The beginning of the increase in the number of new cases of infection (week) | Change (weekly) |
|---|---|---|---|---|---|---|
| Altai Territory | ----- | ---- | ----- | 35 | 36 | 1 |
| Amursk Region | ----- | 18 | ----- | 34 | 35 | 1 |
| Arkhangelsk Region | 14 | 15 | 1 | 34 | 35 | 1 |
| Bryansk Region | 15 | 15 | ------ | 35 | 35 | ----- |
| Vladimir Region | 13 | 14 | 2 | 38 | 40 | 2 |
| Volgograd Region | 15 | 17 | 2 | 36 | 39 | 3 |
| Voronezh Region | 15 | 16 | 1 | 34 | 37 | 3 |
| Ivanovo Region | 15 | 15 | ------ | 38 | 40 | 2 |
| Kaliningrad Region | ----- | 15 | ------ | 36 | 37 | 1 |
| Kaluga Region | 16 | 14 | 2 | 38 | 38 | ----- |
| Karachay-Cherkess Republic | 13 | 15 | 2 | 33 | 33 | ----- |
| Kemerovo Territory | ----- | 22 | ----- | 36 | 38 | 2 |
| Kirov Region | ----- | 21 | ----- | 37 | 40 | 3 |
| Kostroma Region | 14 | 14 | ----- | 39 | 36 | 3 |
| Krasnodar Territory | 14 | 15 | 1 | 36 | 39 | 3 |
| Krasnoyarsk Territory | 14 | 15 | 1 | 38 | 40 | 2 |
| Republic of Crimea | ----- | ----- | ----- | 35 | 35 | ----- |
| Kurgan Region | ----- | 24 | ----- | 38 | 39 | 1 |
| Kursk Region | 15 | 20 | 5 | 38 | 37 | 1 |
| Leningrad Region | 13 | 14 | 1 | 37 | 36 | 1 |
| Magadan Region | ----- | 16 | ----- | 36 | 37 | 1 |
| Moscow | 13 | 14 | 1 | 37 | 39 | 2 |
| Moscow Region | 13 | 8 | 5 | 36 | 40 | 4 |
| Murmansk Region | 13 | 15 | ----- | 36 | 38 | 2 |
| Nenets Autonomous District | 14 | 16 | 2 | 37 | 39 | 2 |
| Nizhny Novgorod Region | 15 | 15 | ----- | 37 | 40 | 3 |
| Novgorod Region | 15 | 16 | 1 | 37 | 39 | 2 |
| Novosibirsk Region | 15 | 16 | 1 | 37 | 39 | 2 |
| Omsk Region | 24 | 24 | ------ | 37 | 36 | 1 |
| Orenburg Region | ----- | 16 | ----- | 35 | 37 | 2 |

| | | | | | | |
|---|---|---|---|---|---|---|
| Oryol Region | 14 | 15 | 1 | 38 | 39 | 1 |
| Penza Region | ----- | 16 | ----- | 34 | 38 | 4 |
| Primorsky Territory | ----- | 16 | ----- | 37 | 41 | 4 |
| Republic of Altai | ------ | 17 | ----- | 37 | 38 | 1 |
| Republic of Kalmykia | ----- | ----- | ----- | ----- | ----- | ----- |
| Republic of Karelia | ----- | ------ | ----- | 37 | 39 | 2 |
| Mari El Republic | 14 | 15 | 1 | 38 | 37 | 1 |
| Republic of Mordovia | 15 | 15 | ----- | 38 | 39 | 1 |
| Republic of Sakha (Yakutia) | 14 | 16 | 2 | 34 | 36 | 2 |
| Republic of North Ossetia-Alania | 13 | 15 | 2 | 40 | 33 | 6 |
| Republic of Tyva | 14 | 19 | 5 | 36 | 38 | 2 |
| Rostov Region | ----- | 16 | ----- | 35 | 36 | 1 |
| Ryazan Region | 15 | 15 | ----- | 37 | 39 | 2 |
| St. Petersburg | 14 | 16 | 2 | 36 | 40 | 4 |
| Saratov Region | ----- | 16 | ----- | 35 | 36 | 1 |
| Sakhalin Region | ----- | ----- | ---- | 37 | 40 | 3 |
| Sverdlovsk Region | ----- | 17 | ----- | 35 | 40 | 5 |
| Sevastopol | ----- | ----- | ----- | ----- | 41 | ----- |
| Smolensk Region | 15 | 16 | 1 | 37 | 39 | 2 |
| Stavropol Territory | 13 | 15 | 2 | 38 | 38 | ----- |
| Tambov Region | 15 | 15 | ----- | 37 | 38 | 1 |
| Tver Region | 14 | 22 | 8 | 14 | 22 | 8 |
| Tomsk Region | ----- | 14 | ----- | 38 | 36 | 2 |
| Tula Region | 14 | 15 | 1 | 37 | 38 | 1 |
| Ulyanovsk Region | 15 | 15 | ----- | 34 | 36 | 2 |
| Khanty-Mansi Autonomous Area | ----- | 16 | ----- | 37 | 36 | 1 |
| Chuvash Republic | 13 | 14 | 1 | 37 | 40 | ----- |
| Yamal-Nenets Autonomous District | ----- | 24 | ----- | 37 | 38 | 1 |
| Yaroslavl Region | 15 | 16 | 1 | 37 | 37 | ----- |
