## Supplementary table 4 for "Online queries as a criterion for evaluation of the epidemiological status and effectiveness of COVID-19 epidemic control measures"

**Supplementary Table 4. Statistics of levofloxacin-related queries in Russia**

| Region | New cases of infection | The number of levofloxacin-related queries | The levofloxacin queries/new infection cases ratio |
|---|---|---|---|
| Moscow | 963941 | 822911 | 0.9 |
| St. Petersburg | 360459 | 285314 | 0.8 |
| Tver Region | 32688 | 40679 | 1.2 |
| Republic of North Ossetia-Alania | 15473 | 26417 | 1.7 |
| Leningrad Region | 34846 | 44392 | 1.3 |
| Arkhangelsk Region | 55642 | 18857 | 0.3 |
| Novgorod Region | 26582 | 17263 | 0.6 |
| Kaliningrad Region | 27421 | 16781 | 0.6 |
| Mari El Republic | 11532 | 13497 | 1.2 |
| Altai Territory | 16090 | 78015 | 4.8 |
| Stavropol Territory | 46642 | 103974 | 2.2 |
| Moscow Region | 963941 | 400554 | 0.4 |
| Nenets Autonomous District | 1001 | 558 | 0.6 |
| Smolensk Region | 25618 | 31091 | 1.2 |
| Bryansk Region | 32158 | 51433 | 1.6 |
| Vladimir Region | 26509 | 62799 | 2.4 |
| Ryazan Region | 23946 | 61161 | 2.6 |
| Nizhny Novgorod Region | 93747 | 162097 | 1.7 |
| Krasnodar Territory | 37301 | 286333 | 7.7 |
| Penza Region | 37018 | 56406 | 1.5 |
| Chuvash Republic | 21014 | 47122 | 2.2 |
| Ulyanovsk Region | 46895 | 66214 | 1.4 |
| Kaluga Region | 35567 | 36908 | 1.0 |

| | | | |
|---|---|---|---|
| Kaluga Region | 28984 | 54205 | 1.9 |
| Volgograd Region | 47980 | 104269 | 2.2 |
| Kostroma Region | 18120 | 16456 | 0.9 |
| Tambov Region | 25406 | 56262 | 2.2 |
| Oryol Region | 29623 | 28292 | 1.0 |
| Tula Region | 32121 | 61625 | 1.9 |
| Yaroslavl Region | 31751 | 39911 | 1.3 |
| Ivanovo Region | 29864 | 28996 | 1.0 |
| Kursk Region | 29731 | 78557 | 2.6 |
| Khanty-Mansi Autonomous Area | 50072 | 34701 | 0.7 |
| Saratov Region | 48057 | 149431 | 3.1 |
| Kemerovo Region | 31169 | 82028 | 2.6 |
| Orenburg Region | 36931 | 79134 | 2.1 |
| Amursk Region | 20688 | 16179 | 0.8 |
| Voronezh Region | 65749 | 122168 | 1.9 |
| Primorsky Territory | 39688 | 34037 | 0.9 |
| Sverdlovsk Region | 76315 | 145440 | 1.9 |
| Tomsk Region | 29879 | 18982 | 0.6 |
| Republic of Mordovia | 17448 | 32854 | 1.9 |
| Krasnoyarsk Territory | 60660 | 286333 | 4.7 |
| Novosibirsk Region | 34917 | 176262 | 5.0 |
| Kurgan Region | 17112 | 23857 | 1.4 |
